# Modelling, prediction and design of national COVID-19 lockdowns by stringency and duration

**DOI:** 10.1101/2021.03.12.21253454

**Authors:** Alberto Mellone, Zilong Gong, Giordano Scarciotti

**Affiliations:** Department of Electrical and Electronic Engineering, Imperial College London, London, SW7 2AZ, UK

## Abstract

The implementation of lockdowns has been a key policy to curb the spread of COVID-19 and to keep under control the number of infections. However, quantitatively predicting in advance the effects of lockdowns based on their stringency and duration is a complex task, in turn making it difficult for governments to design effective strategies to stop the disease. Leveraging a novel mathematical “hybrid” approach, we propose a new epidemic model that is able to predict the future number of active cases and deaths when lockdowns with different stringency levels or durations are enforced. The key observation is that lockdown-induced modifications of social habits may not be captured by traditional mean-field compartmental models because these models assume uniformity of social interactions among the population, which fails during lockdown. Our model is able to capture the abrupt social habit changes caused by lockdowns. The results are validated on the data of Israel and Germany by predicting past lockdowns and providing predictions in alternative lockdown scenarios (different stringency and duration). The findings show that our model can effectively support the design of lockdown strategies by stringency and duration, and quantitatively forecast the course of the epidemic during lockdown.

## Introduction

By the 2nd of January 2020, forty-one hospital cases of infection by the severe acute respiratory syndrome coronavirus 2 (SARS-CoV-2) were confirmed in Wuhan^1^. By the 11th of March 2020, the coronavirus disease 2019 (COVID-19) that SARS-CoV-2 causes in humans was declared a global pandemic^2^. A relatively high infectivity^3, 4^, combined with a substantial fraction of the cases requiring hospital admission and intensive care^5, 6^, makes COVID-19 a threat to the public health. In the absence of effective treatments, capable of preventing a severe and potentially fatal evolution of the disease in humans, and while a vaccine is not widely available, governments around the world are implementing social distancing measures to curb the proliferation of the epidemic^7^, thus containing the number of complications and deaths. The most drastic of these measures is the implementation of nation-wide lockdowns. National lockdowns might differ in how restrictive they are, but all share the same goal: constraining the majority of the population to stay at home, avoiding travel and meeting people outside of one’s own household. Lockdowns have successfully curbed the spread of the disease^8, 9^ and prevented national health services from being overwhelmed by an otherwise unbearably large number of severe infections. Even though since December 2020 vaccination campaigns have started^10^ in several countries, it is still unclear when the roll-out will guarantee that a sufficient percentage of the population is vaccinated, as well as how effective the jab will be in preventing complications arising from new coronavirus variants^11^. This has made governments cautious about lifting social distancing policies, which as of March 2021 are still in place in the vast majority of the countries.

Mathematical models of epidemics play a crucial role in tackling the proliferation of the disease. On the one hand, models allow for predictions of how the disease will spread in the future, thus providing an estimate, for instance, of the number of people who will become ill (and to what extent), of the number of patients who will recover and of the number of casualties resulting from the epidemic. On the other hand, models are fundamental in driving governmental plans to curb the spread of the disease, such as restrictions on people’s movements, mass testing strategies and, when a vaccine is available, mass vaccination campaigns. The most popular models are the mean-field compartmental models. The population is split in multiple, mutually exclusive compartments. Each of these compartments represents a stage of the disease, with individuals moving from a compartment to another as the disease progresses. Several compartmental models have been developed for COVID-19. Cooper et al. designed^12^ a SIR model to investigate the spread of the disease in a number of communities, whereas Kyrychko et al. modelled^13^ the outbreak in Ukraine in a SEIR-type fashion. Kucharski et al. considered^14^ a stochastic SEIR model to study the spread of infections in and originating from Wuhan, while Davies et al. investigated^15^ the UK epidemic using a stochastic SEIR-like model stratified by age groups. Giordano et al. proposed^16^ a compartmental model in which the infected individuals are differentiated both by disease severity and diagnosis stage. Ndaïrou et al. introduced^17^ a compartmental model with a focus on super-spreader individuals. Della Rossa et al. investigated^18^ the impact of Italian inter-regional infections by modelling the whole country population as a network of compartmental models. As far as the effects of radical non-pharmaceutical interventions such as lockdowns is concerned, Flaxman et al. proposed^19^ a model that estimates, backwards from observed deaths, the disease transmission and studied the implications of lockdown timing. Oraby et al. considered^20^ a continuous-time Markov chain compartmental model and focused on the effect of lockdowns on the hospitalisations. Other mean-field models^21, 22^ explicitly take into account the lockdown effects by changing the model structure when lockdowns are introduced or lifted, but the approaches remain compartmental.

## Results

### Model

A core, simplifying, assumption of mean-field compartmental models is that all individuals in the population have the same degree of interaction with everyone else, with the epidemic spreading as a result of contacts between infectious and susceptible individuals. This assumption is violated when lockdown measures are introduced nation-wide. At the start of a lockdown the population is split in two categories that are characterised by remarkably different levels of social interaction. A part of the population, usually a minority, either is partially exempt from lockdown restrictions (for instance pupils when schools are open, healthcare workers or other essential workers) or does not abide by them. Therefore, this category of individuals keeps having a high level of social interaction. The rest of the population isolates at home within their own household, thus limiting their social interactions to a very reduced number of other individuals. Contacts with people outside of one’s own household are still possible (for instance when visiting an essential shop, or when attending healthcare appointments), but they are generally rare. Moreover, people in households cannot even be considered as one compartment, but rather as a multitude of small units which evolve independently from one another. In other words, on the day in which a lockdown is enforced, the population undergoes an abrupt change in its dynamic behaviour, which undermines the assumption on which traditional mean-field compartmental models are grounded. The FL-Hybrid epidemic model that we propose in this paper overcomes this fundamental limitation, in that it is able to differentiate between these two categories of individuals characterised by different levels of social interaction. The FL-Hybrid model exploits a hybrid mathematical framework^23^ that allows modelling the interaction between discrete events (*e*.*g*. governments introducing lockdowns) on continuous-time dynamics (*e*.*g*. compartmental models).

The FL-Hybrid model switches between two fundamental phases: the free phase and the lockdown phase. If the population is allowed to move freely (for example at the start of the epidemic or after a lockdown is lifted) the model is in the free phase. In this phase the assumption of traditional compartmental models is verified, therefore the free phase is described by a sub-model that is a variation of a standard SIR model. We refer to this classical model as SUDER, in which the population is partitioned in five disease stages: S, susceptible; U, undetected or undiagnosed infected; D, detected or diagnosed infected; E, extinct (dead); R, recovered. When a lockdown is enforced by the government as a result of a dramatic increase in infections, the model switches to the lockdown phase. In this phase a minority of the population (including, *e*.*g*., essential workers) is still assumed to move freely (for the most part), thus its behaviour is still accurately described by the SUDER sub-model. However, the majority of the individuals are forced to avoid unnecessary contacts with anyone outside of their household, thus determining a different evolution of the epidemic within this portion of the population. The dynamics of the epidemics among these individuals is described by the HP, *i*.*e*. Household Partitions, sub-model. In the HP sub-model the household, rather than the individual, is the fundamental unit and represents a small group of individuals who often come in contact with each other, but rarely interacts with the rest of the population. Hence, a household with zero infected individuals at the start of the lockdown will most likely keep this status until the end of the lockdown. Susceptible members in a household might still be infected from external contacts, but the probability of this happening is low. Instead, if one or multiple infected individuals are present in the household, then the HP sub-model describes the spread of the disease among the household members, who then, if infected, go through the usual U, D, E, and R stages of infection. The relevance of the FL-Hybrid model we introduce is not only due to addressing mathematically the abrupt change of social behaviour caused by a lockdown. The importance of this new model also lies in the fact that it provides the policymakers with a tool to assess the impact of a past lockdown on the course of the epidemic as well as to plan for potentially new lockdowns, should the cases be on a sharp rise in the future. In fact, a parameter that the model allows tuning is the *stringency* of the lockdown, *i*.*e*. the percentage of population effectively constrained in their own household. Different stringency levels are reflected in real life by, for instance, what type and how many workers are considered essential, thus being allowed to move freely, but also how strictly the rules are enforced and, therefore, how many individuals are expected to abide by them. A second parameter that the model allows tuning is the *duration* of the lockdown, which may be crucial to evaluate the optimal time to lift it, thus avoiding a new surge in cases as well as preventing an overly prolonged paralysis of the economy. Apart from stringency and duration, which are parameters for the policymakers to design, the rest of the model parameters can be estimated from analysis of the available data, specifically the number of active COVID-19 infections in a given country, the cumulative number of deaths, and the cumulative number of recoveries. The interactions among the free phase and lockdown phase and, within these, among the sub-models and their inner compartments are shown in Figure 1. A detailed discussion of the model considerations, assumptions and parameters is provided in the Methods.

**Figure 1.**
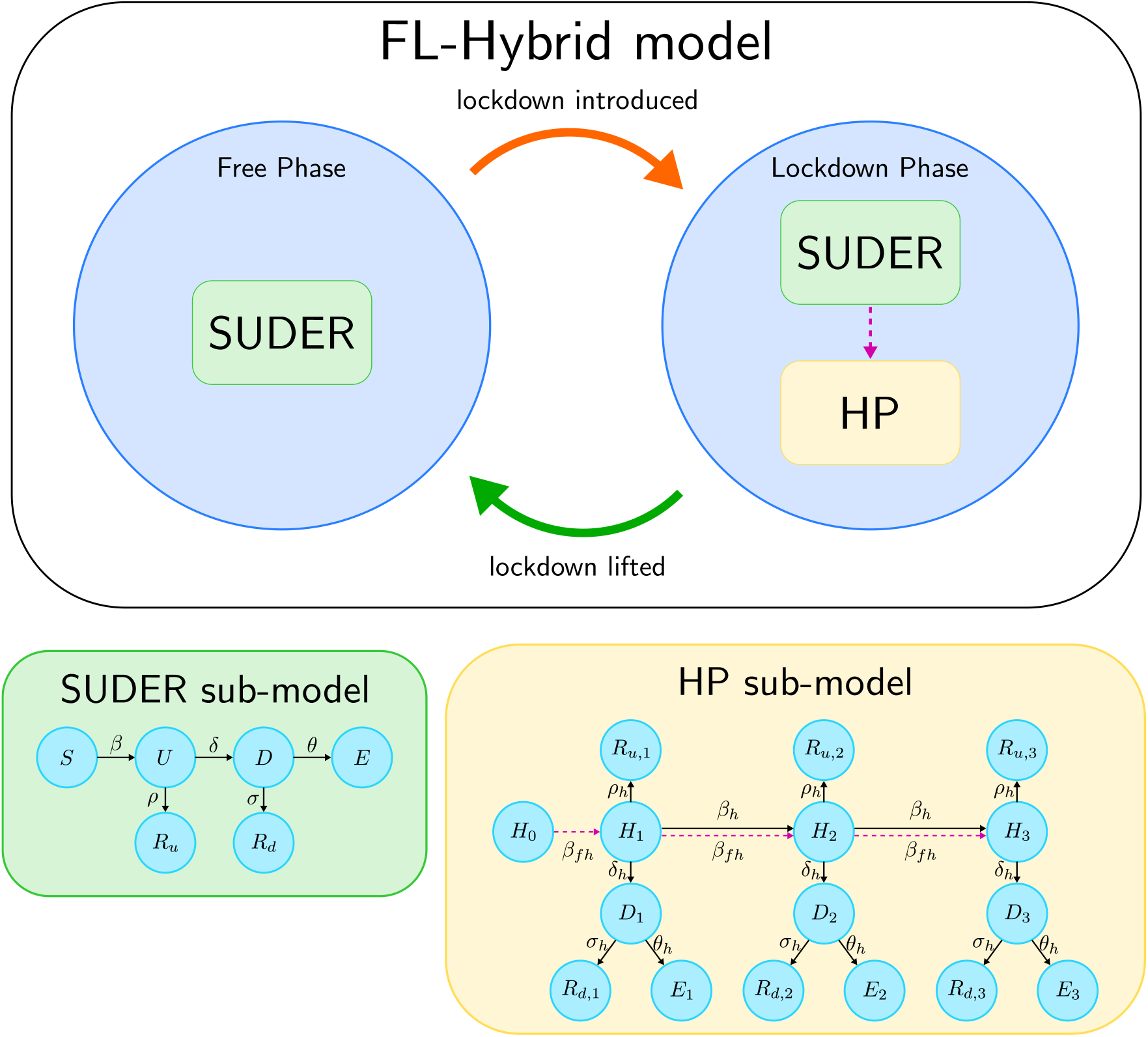
The FL-Hybrid model. Graphical scheme illustrating the switching between the free and lockdown phases of the FL-Hybrid (Free-to-Lockdown Hybrid) model, as well as the interactions between the sub-models operating within them. The SUDER sub-model describes the epidemic dynamics within the whole population in the free phase and within the fraction of the population not isolating during a lockdown. The HP sub-model describes the epidemic dynamics within the fraction of population isolating in their own household during a lockdown. The dashed magenta line represents the potential introduction of infections in isolating households from rare but unavoidable external contacts with individuals in the SUDER-sub-model.

### Findings and discussion

To illustrate the effectiveness of our model we present two case studies, namely the evolution of the COVID-19 epidemics in Israel and Germany. The reason why these countries have been selected as an illustrative example is twofold: on the one hand, a full set of data, including recoveries, is provided and updated daily; on the other hand, these two countries enforced at least two nation-wide lockdowns at different times, so their data allow us to illustrate our model’s ability to predict the epidemic evolution during a successive lockdown based on data from the previous ones.

### Stringency

We first illustrate the effect of changing the strength of lockdown policies on the number of active cases and deaths. We define the active cases of infection at any given time as the difference between the cumulative number of diagnosed infections and the sum of cumulative deaths and recoveries. In our model the active cases are represented by the people in the D (diagnosed) compartment, whereas the deaths are represented by the population in the E (extinct) compartment. Figures 2a and 2b show the change in the number of active cases and deaths as a result of changing the stringency of a lockdown in Israel. Active cases and deaths are reported as fraction of the entire population. We focus on the nation-wide lockdown imposed for thirty days, between 18/09/2020 and 18/10/2020 (days 183 and 213 since the start of the epidemic). In both figures, the dashed blue line represents the actual evolution of the detected active cases and deaths in the country as reported by governmental sources (see Data availability). The peak of active cases was recorded at about 0.7901% of the population and after the thirty-day period the active cases dropped to about 0.2860%. At the end of the lockdown the cumulative deaths due to COVID-19 were reported as 0.0249% of the population. The solid blue line represents our model prediction when the percentage of people isolating with their own household is 65%. As the solid and dashed blue lines show very comparable trends, the data is compatible with our model prediction that about two thirds of the population stayed at home. Thus, this fraction becomes our baseline for assessing different lockdown stringency levels (see Methods for further details on the meaning of this percentage). The cyan and magenta lines show our model predictions for the number of detected active cases and deaths if the lockdown had been implemented more or less stringently. Namely, we set the percentage of people isolating at home to 80% and 50% respectively. As shown in Figure 2a a more stringent lockdown would have resulted in an earlier and lower peak of active infections, with slightly more than 0.6478% of the population representing active cases of infection at the peak and about 0.2230% at the end of the lockdown. A less stringent lockdown would have resulted in a peak of active cases of about 0.9413% of the population, dropping to about 0.4671% at the end of the lockdown. The number of deaths as predicted by our model and illustrated in Figure 2b show that if the lockdown had been more stringent, the number of deaths at the end of the lockdown would have been about 0.0228% of the population, whereas a milder lockdown would have caused about 0.0272% of the population to die from COVID-19 by the end of the lockdown. Finally, the red line in both figures portraits the scenario in the case that no lockdown had been enforced. Both the active cases and the deaths experience a dramatic surge, which does not show any sign of attenuation after a thirty day period. Figures 2c and 2d show an analogous behaviour when a change in lockdown stringency is considered in relation to the national lockdown imposed in Germany between 23/03/2020 and 12/05/2020 (days 23 to 73 since the beginning of the outbreak), lasting fifty days. Our model suggests that the government data (see Data availability) on active cases and deaths (dashed blue lines in the figures) are compatible with 80% of the population isolating within their own household. This is evident by our model prediction, which replicates (solid blue line) the real trends when such a percentage of population is assumed to isolate at home. This also represents our baseline for comparisons with more or less stringent lockdowns. As shown in Figure 2c, this scenario yields a peak of about 0.0877% of the population being active cases, which the lockdown managed to reduce to 0.0219% after the fifty-day period. Figure 2d shows that the deaths at the end of the lockdown represent about the 0.0093% of the population. As before, the cyan and magenta lines illustrate the scenarios of more stringent (90%) and less stringent (60%) lockdowns. In Figure 2c the former produces a lower and flatter curve of active cases, peaking at about 0.0566%, while the latter results in a higher peak at 0.1648%, just slightly under double the actually diagnosed cases. At the end of the milder lockdown the number of active cases, although declining, is similar to the number of active cases at peak when an 80% stringent lockdown is implemented. Figure 2d illustrates that, as expected, the number of deaths at the end of the lockdown increases as the lockdown stringency decreases, with the final deaths being 0.0053% of the population with a 90% stringency and 0.0251% of the population with a 60% stringency. Finally, the red lines in both Figures 2c and 2d show that imposing no lockdown (0% stringency) would have caused a dramatic rise in both diagnosed cases and deaths. From Figure 2 our model suggests that the lockdown is an extremely effective way to curb the spread of the infection among the population, thus confirming previous findings^7, 9^. Additionally, our model allows making two types of assessment. Firstly, it is possible to infer how large the proportion of population actually isolating is. This enables policymakers to evaluate, for example, whether the isolation rules are enforced in an adequate way or if the number of workers considered essential can be increased or must be reduced. Secondly, our model helps to quantify the lockdown effectiveness by predicting potential outcomes, in terms of number of active cases and deaths, when a lockdown is implemented with different levels of strength. Figure 2 shows that even milder forms of lockdown (*i*.*e*. making sure that about half of the population is isolating) help to make the curve of active infections flatter, compared to the case of no restrictions, thus avoiding an extremely large number of infections and deaths.

**Figure 2.**
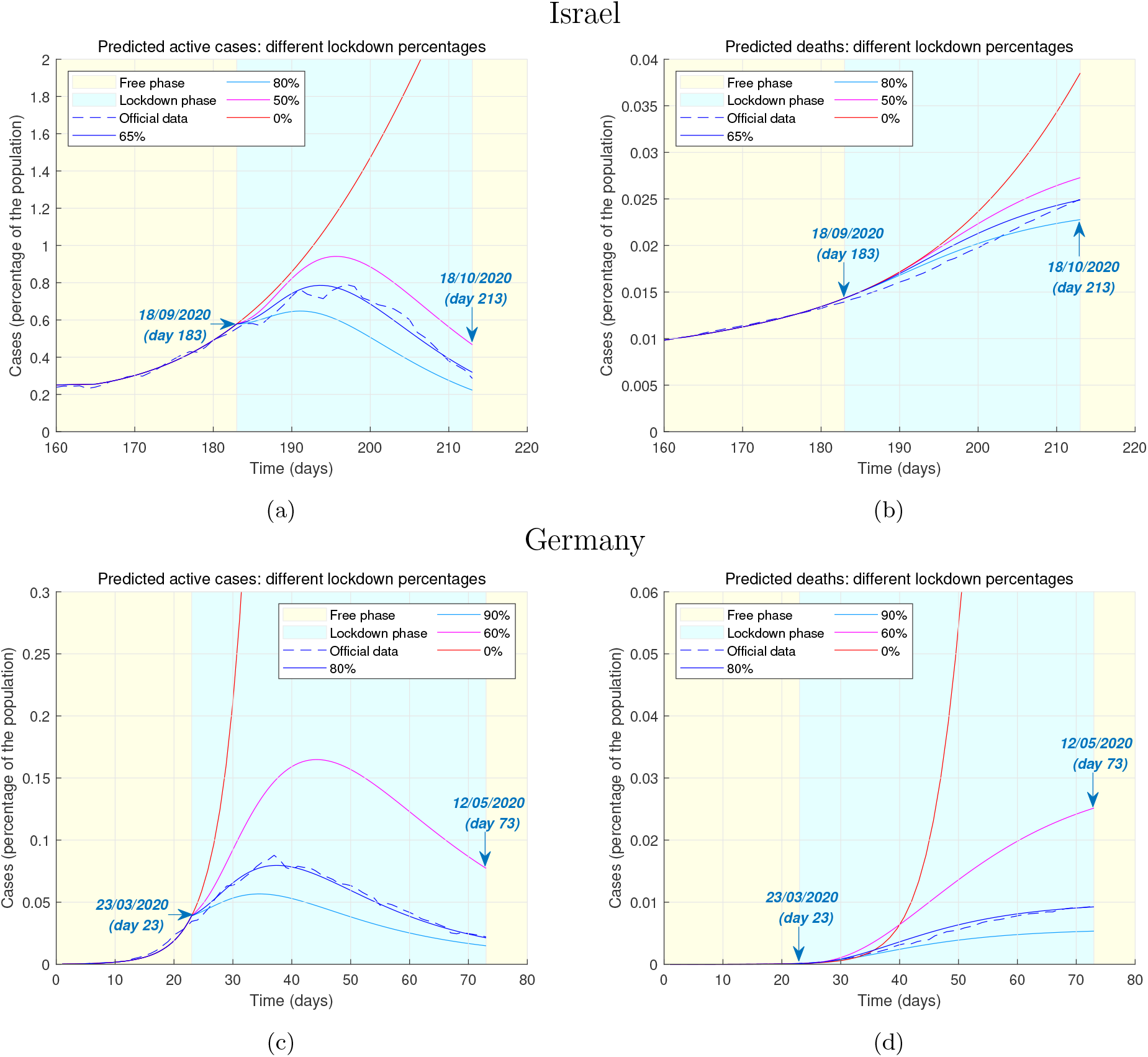
Predicted epidemic evolution under different lockdown stringency. Evolution of the COVID-19 epidemic in Israel and Germany as predicted by our model, based on available government data. **2a**, Actual evolution and model predictions of the active cases of infections in Israel by lockdown stringency. **2b**, Actual evolution and model predictions of the deaths in Israel by lockdown stringency. **2c**, Actual evolution and model predictions of the active cases of infections in Germany by lockdown stringency. **2d**, Actual evolution and model predictions of the deaths in Germany by lockdown stringency.

### Duration

Having assessed the effects of lockdown stringency on predicted number of infections and deaths, we now consider the implications of different lockdown durations. Figures 3a and 3b illustrates this for the case study of the lockdown imposed in Israel between 18/09/2020 and 18/10/2020. As before, the dashed blue line represents the government data, whereas the solid blue line represents the model prediction with a lockdown stringency baseline of 65% (as discussed above). While the lockdown in Israel lasted thirty days in this instance, we consider alternative scenarios in which a lockdown with the same stringency is lifted later or earlier, and we assess our model predictions on a window of thirty days after the lockdown end. The cyan line represents the evolution of active cases and deaths when the lockdown lasts fifty days, while the red line illustrates the same quantities for a lockdown lasting twenty days. Figure 3a shows that a longer lockdown would have been able to steer the number of active cases to very low figures, whereas a slightly shorter lockdown would have caused a dramatic slowdown in the fall of active cases, signalling a resurgence of the new infections. The benefits of longer lockdowns are also evident in Figure 3b, in which the fifty-day lockdown produces a stronger halt in the rise of number of deaths, compared to the thirty-day one actually enforced. On the contrary our model predicts that deaths would have further risen considerably after the lockdown, had this been lifted after just twenty days. In Figures 3c and 3d we make a similar assessment for the German nation-wide lockdown between 23/03/2020 and 12/05/2020, considering hypothetical scenarios in which the lockdown would have lasted forty or seventy days, instead of the actual fifty days, and predicting the course of the epidemic in a twenty-day window after the lockdown. The stringency is set at the baseline of 80%. While the forty-day lockdown scenarios produce trends similar to the Israeli case, in that lifting a lockdown slightly earlier would have produced a substantial halt in the drop of number of active cases and, consequently, in the rise of the number of deaths, it is interesting to observe that prolonging the lockdown for twenty more days would not have had as considerable benefits as if Israel had increased the duration of its lockdown. The number of active cases does experience a further drop when the lockdown is longer, while the rise of the number of deaths does appear to slow down. However, these positive outcomes are not as considerable as those produced by a longer lockdown in Israel (which was shorter to begin with). Ultimately, our model suggests that a lockdown duration of about fifty days yields the most benefits, as confirmed by the fact that, had Israel enforced this duration, the additional decrease of active cases and, consequently, of deaths would have been significant. Longer lockdowns might not yield substantial improvements, yet they might be deeply damaging from a social or economic point of view.

**Figure 3.**
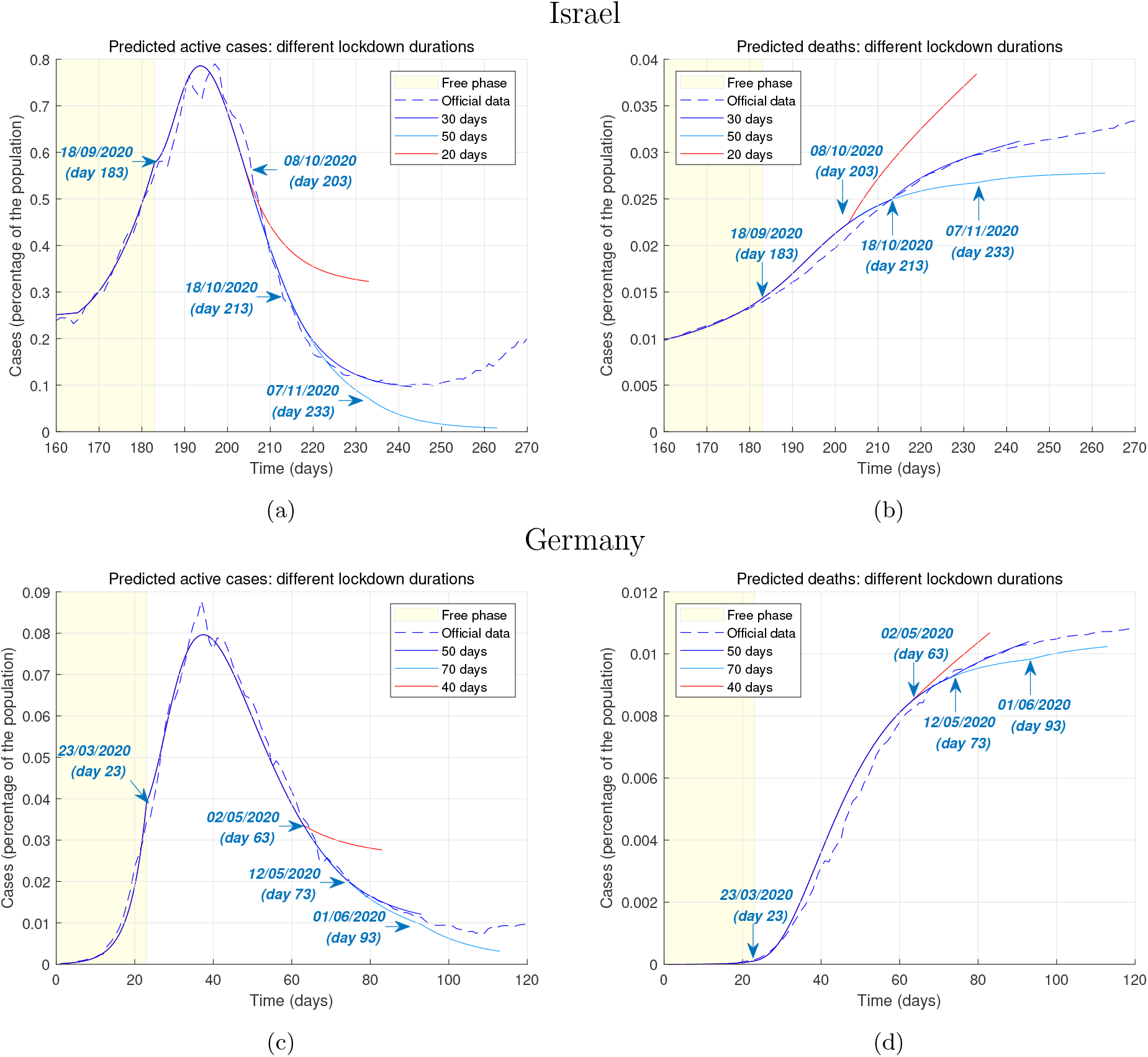
Predicted epidemic evolution under different lockdwon durations. Evolution of the COVID-19 epidemic in Israel and Germany as predicted by our model, based on available government data. **3a**, Actual evolution and model predictions of the active cases of infections in Israel by lockdown duration. **3b**, Actual evolution and model predictions of the deaths in Israel by lockdown duration. **3c**, Actual evolution and model predictions of the active cases of infections in Germany by lockdown duration. **3d**, Actual evolution and model predictions of the deaths in Germany by lockdown duration.

### Subsequent lockdown

At last, Figure 4 demonstrates that our model is able to predict the evolution of a new lockdown only based on data from previous lockdowns. This kind of prediction is different from the previous ones we presented, in that we no longer provide alternative scenarios in which different types of lockdown are implemented. Instead, here we suppose to be at the start of the second lockdown, and we assume that no data is available from the future course of the epidemic. The model is able to reasonably predict the number of active cases and infections in a future time window by leveraging the data collected in the first lockdown. This is a powerful tool that enables policymakers to design the lockdown in order to achieve, for instance, a desired peak of the active cases or to limit the total number of deaths. In Figures 4a and 4b we consider the second Israeli national lockdown starting on 27/12/2020 (day 283 of the epidemic). In our model we set a 60% lockdown stringency - instead of the first lockdown’s 65% - to predict the course of the epidemic in the first fifteen days of the second lockdown. This is compatible with the fact that in the first days this lockdown was milder than the previous one^24^. Based on data from the previous lockdown, our model is able to predict that the future peak of the active cases of infections would be about 0.9% of the population. The future increase of the total number of deaths is also accurately reproduced. Figures 4c and 4d illustrate analogous findings for the second lockdown in Germany, starting on 02/11/2020 (day 247 of the epidemic). This second lockdown is less stringent than the first one^24^, which is compatible with a 70% stringency in our model, instead of the first lockdown’s 80%. Moreover, the drastically increased testing capacity also reduced the case fatality rate of coronavirus^25^. Using data collected in the first lockdown and adjusting for a smaller case fatality rate, we predict the course of the epidemic for the first fifteen days of lockdown. Our model successfully predicts that the peak of the active cases is approximately at 0.37% of the total population, and is able to provide consistent forecasts of the future trends of deaths. It is important to remark that these predictions of the future course of the epidemic are to be considered an initial and provisional guess of the future trends. When real data is available, this should be used to assess the effectiveness of the lockdown and, by feeding it back to the model, produce improved future predictions. The motivations, findings and implications of our model are summarised in Table 1.

**Table 1:** Policy summary.

|  |  |
| --- | --- |
| Background | The introduction of a national lockdown is an effective mean of containing the spread of COVID-19. The implementation of a lockdown requires a careful design aimed at limiting the number of infections and, consequently, deaths. We propose a model that is able to support this process by providing future predictions on the course of the epidemic for different stringency levels and durations of the lockdown. |
| Main findings and limitations | **Stringency.** Even mild lockdowns, *i.e.* isolating about 50% of the population, result in dramatic improvements compared to not implementing any form of lockdown. However, ensuring that around 80% of the population isolate within their household yields reasonably better performances, whilst more aggressive (*i.e.* more than 80% isolating population) lockdowns produce negligible additional benefits, which are potentially outbalanced by the financial and social drawbacks caused by the restrictions.<br>**Duration.** Lifting a lockdown too early (*e.g.* before 20 days since it started) is likely to induce a sharp new increase of infections and deaths or, at best, to cause a halt in the decrease of cases. A lockdown duration of around 50 days produces substantial improvements in terms of limiting the number of infections and deaths. While enforcing a lockdown for longer periods (*e.g.* more than 50 days since it started) has been found to yield marginal improvements, in the long run the negative social and business implications of prolonged restrictions might outweigh such advantages.<br>**Prediction.** By leveraging data collected in a previous lockdown, it is possible to accurately predict the peak of active cases and deaths when a new lockdown is enforced. As customary in modelling studies, our model-based predictions are based on reasonable assumptions, but the real evolution of the epidemic depends on how prompt and effective the actual public health and safety policies are. |
| Policy implications | Our model can support policymakers to plan ahead of a lockdown by providing them with a tool to predict the course of the epidemic as a result of different lockdown stringency levels and/or durations. This is paramount to achieve a trade-off between the negative impact on business and society and the crucial goal of limiting infections and deaths. |

**Figure 4.**
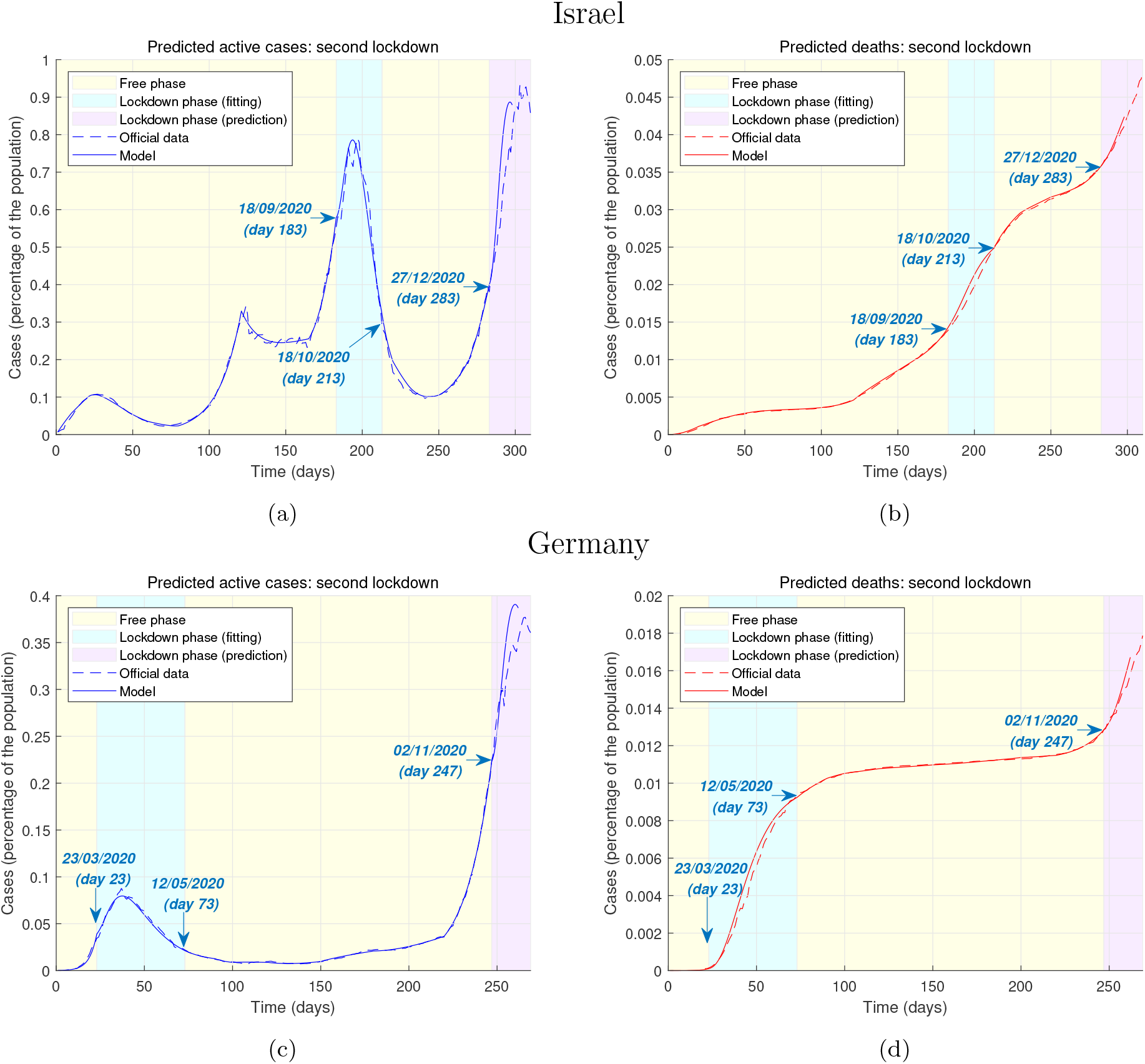
Predicted epidemic evolution during the second lockdown. Evolution of the COVID-19 epidemic in Israel and Germany during a second national lockdowns as predicted by our model, based on available government data. The prediction of the evolution during the second lockdown (enforced at the start of the second “wave”) is performed assuming no data from the future course of the epidemic is available at the start of the lockdown. **3a**, Actual evolution and model prediction of the active cases of infections in Israel throughout the epidemic. **3b**, Actual evolution and model predictions of the deaths in Israel throughout the epidemic. **3c**, Actual evolution and model predictions of the active cases of infections in Germany throughout the epidemic. **3d**, Actual evolution and model predictions of the deaths in Germany throughout the epidemic.

## Methods

### FL-Hybrid model

The FL-Hybrid (Free-to-Lockdown Hybrid) model is a hybrid dynamical model^23^ switching between two modes, which we call phases: the *free phase* and the *lockdown phase*. Both the free phase and the lockdown phase of the model are described by sets of ordinary differential equations and represent the so-called flow of the hybrid model. The switching action (*i*.*e*. the so-called jump set of the hybrid model) corresponds to the government’s action of enforcing or lifting a lockdown.

The free phase models the evolution of the epidemic when the entire population is allowed to move freely and no strict policy requiring the individuals to stay at home and avoid contacts with others is enforced. This phase typically occurs at the beginning of the epidemic or after a period of lockdown, when social distancing measures are relaxed as a result of a drop in number of diagnosed infections.

The lockdown phase models the evolution of the epidemic when a lockdown is imposed. The population is divided into two categories, which we call the *free population* and the *lockdown population*. The free population is a minority that maintains a high number of interactions with other individuals. For example, this is the case of key workers, who are partially exempt from isolation to carry out essential jobs, but also takes into account that some individuals do not comply with the regulations. On the other hand, the lockdown population is assumed to have a drastically reduced number of social interactions. This population is split into *households*, which are for simplicity assumed to be of fixed size of three members (the rationale of this number is explained later).

We now describe the mathematical model in more detail and we discuss the modelling assumptions later.

### Free phase

In the free phase the epidemic evolves according to a standard compartmental sub-model which we call SUDER sub-model. The SUDER sub-model is a dynamical model consisting of six ordinary differential equations. Each equation characterises the change over time of the proportions of population experiencing a specific stage of the disease. This model describes the dynamics of the epidemic when no lockdown measures are introduced. The system of equations is given by

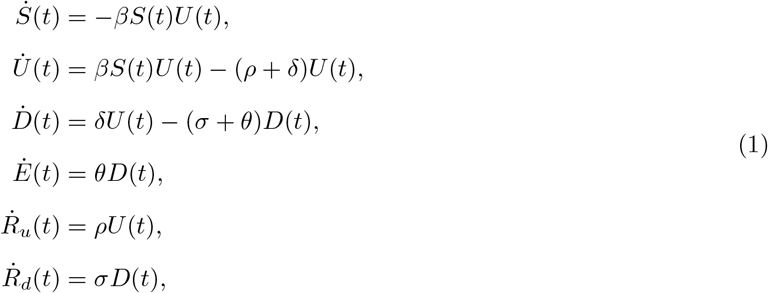

where *S* (Susceptible), *U* (infected Undetected), *D* (infected Detected), *E* (Extinct), *R*_*u*_ (undetected Recovered), and *R*_*d*_ (detected Recovered) are the proportions of the population at each stage of the disease, while the Greek letters represent the parameters of the model and are positive numbers. In particular,

- *β* is the disease transmission rate from an undetected infected person to a susceptible person. This is the probability that an undetected person transmits the infection to a susceptible person multiplied by the average number of contacts per person. This parameter is dependent both on the infectivity of the disease and on the number of close contacts between individuals. Therefore this parameter is reduced when social distancing measures are implemented.
- *δ* is the probability rate of detection, *i*.*e*. the probability that an undetected infected person becomes detected after any form of diagnosis. This parameter increases as the scale and efficiency of mass testing and contact tracing policies are improved.
- *ρ* and *σ* are the probability rates of recovery of undetected and detected infected people, respectively. Undetected people are generally asymptomatic or develop very mild symptoms, compared to detected people who might develop life-threatening conditions. Therefore, *ρ* is generally higher than *σ*.
- *θ* is the mortality rate of detected people and is lowered by more effective treatments.

### Lockdown phase

In the lockdown phase the epidemic evolves according to the interaction of two sub-models, one for the free population and one for the lockdown population. This interaction results in a system of twenty-two ordinary differential equations.

Among the free population the epidemic evolves according to the SUDER sub-model, therefore six out of twenty-two equations are analogous to the ones previously introduced and are given by

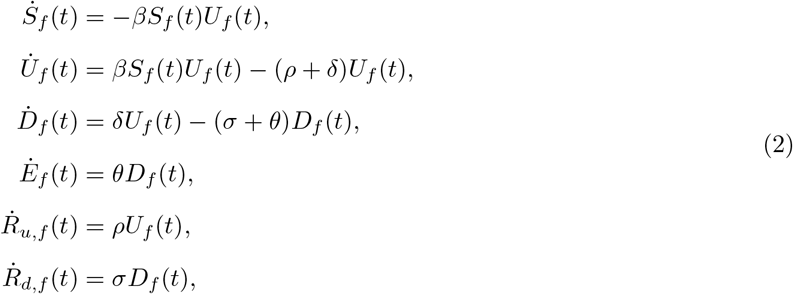

where both the Latin and Greek letters have the same interpretations as in (1) and the subscript *f* specifies that the quantities characterise the free population while a lockdown is enforced.

The remaining sixteen equations describe the evolution of the epidemic among the lockdown population and constitute the so-called HP (for Household Partitions) sub-model. This sub-model aims at considerably simplifying the complex dynamics of people isolating in lockdown, while at the same time capturing the fundamental behaviour of the progression of the disease. The core of the HP sub-model is the household, which is a unit composed of 3 individuals. The assumption on the fixed number of individuals is justified later. The equations model the spread of the disease among members of the same household who are observing the lockdown measure, but also the fact that households do not isolate perfectly and new infections can be introduced in a household when its members get in contact with infected people from the free population. Let *H*_*i*_ denote the number of households with *i* undetected infected, with *i* = 0, 1, 2, 3. Then the equations describing their dynamics are given by

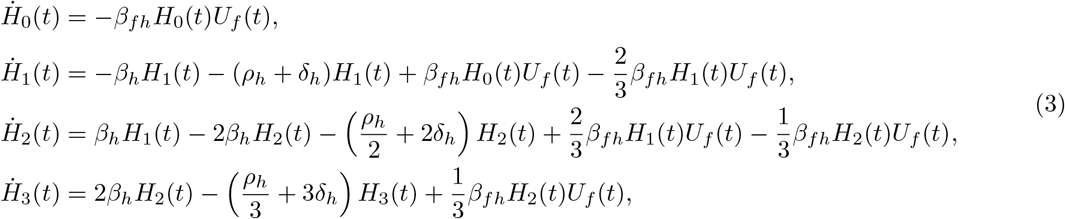

where

- *β*_*h*_ denotes the probability rate that an infected household member infects another susceptible person within the household. This parameter depends on the level of interaction between members of the same household, and increases as household members interact more.
- *ρ*_*h*_ denotes the probability rate that an undetected household member recovers before infecting other people within the household. Again, this depends on the level of household members’ interaction, but decreases as members interact more.
- *δ*_*h*_ denotes the probability rate that an undetected household member becomes detected before infecting other members. This rate depends on the efficiency of testing policies.
- *β*_*fh*_ denotes the disease transmission rate from an undetected person in the free population to a susceptible person in a infection-free household. This parameter depends on the level of exposure of susceptible members in a household to the free population, and decreases as social distancing measures are tightened.

Let *T* be the total population. We now define the quantities *U*_*i*_ = *iH*_*i*_*/T* for *i* = 1, 2, 3, which represent the portions of undetected infected people living in households of type *H*_*i*_. Analogously, we define the quantities *D*_*i*_, *E*_*i*_, *R*_*u,i*_ and *R*_*d,i*_. The set of sixteen equations representing the HP sub-model is then given by

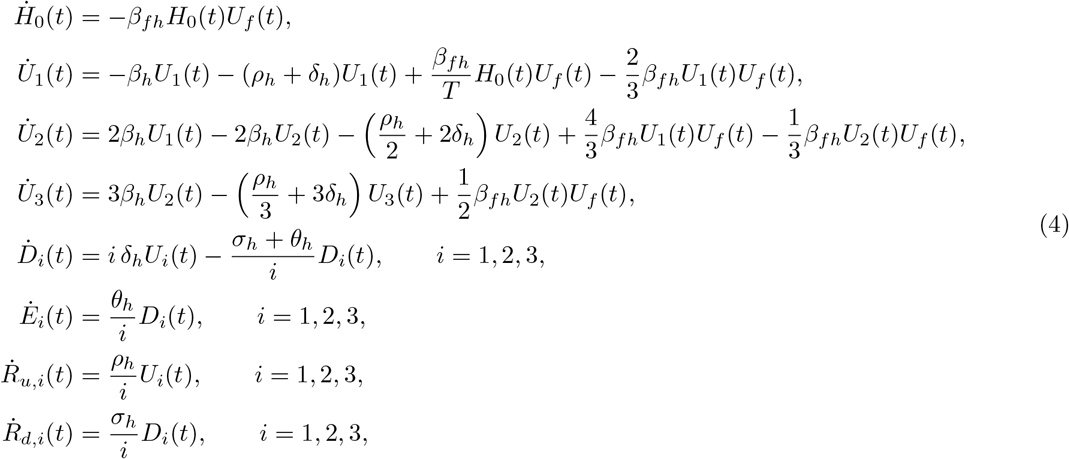

where

- *σ*_*h*_ denotes the probability rate of recovery of a detected infected household member.
- *θ*_*h*_ denotes the mortality rate of a detected infected household member.

Equations (2) and (4) together describe the lockdown phase of the FL-Hybrid model.

The parameters of the model, both in its free and lockdown phases, are estimated using the official data provided by the national health authorities. In particular, we consider the time histories of the total number of COVID-19 diagnosed cases, deaths and recoveries. We then obtain the time history of the COVID-19 active cases by subtracting the deaths and recoveries from the diagnosed cases. The model parameters are then chosen so as to minimise the weighted mean squared errors between the model-predicted time histories of the active cases, deaths and recoveries and the real ones. The model parameters can be periodically updated to reflect the changes in factors like the stringency of social distancing measures, the effectiveness of testing and contact tracing regimes and the efficacy of treatments. These changes might alter the infection, detection, mortality and recovery rates, thus justifying this parameter updating procedure.

### Switching between phases

When the FL-Hybrid model switches between the free phase and the lockdown phase (and vice versa), some conditions on the variables need to be enforced in order to guarantee consistency.

When a lockdown starts, the FL-Hybrid model switches from the free phase to the lockdown phase. To guarantee consistency, the population that is split among the SUDER sub-model compartments in the free phase needs to be distributed among the compartments of the lockdown phase. To do so, we first define the *lockdown percentage L*, which is the percentage of the susceptible and undetected population which will become the lockdown population. Denote by 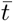 the moment at which the switching between free phase and lockdown phase occurs. Then at the switching the lockdown population will be 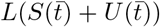. Note that we exclude that detected, extinct or recovered people are part of the lockdown population. This assumption makes the analysis of the model simpler without compromising its accuracy. In fact, the population in *D* is assumed to be perfectly isolated and the population in *E, R*_*u*_, and *R*_*d*_ is not infectious anymore. Therefore these variables do not play an active role in the dynamics of the epidemic, although they still play a fundamental role in tracking the impact of the disease. Distributing these portions of population among the households would have very little impact on the evolution of the epidemic among the lockdown population, and for simplicity they can be considered part of the free population. The free population is therefore 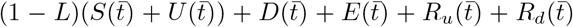. Consequently, the initial conditions for the SUDER sub-model of the free population in the lockdown phase are 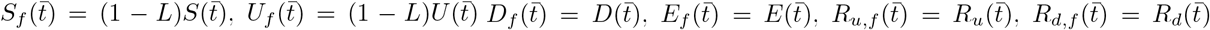. As far as the lockdown population is concerned, this has to be split appropriately into households. The total number of households is given by

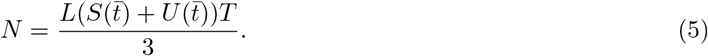

Let *a*_*i*_ be the proportion of households with *i* undetected infected at the beginning of the lockdown, *i*.*e*.

*H*_*i*_ = *a*_*i*_*N*, for *i* = 0, 1, 2, 3. The initial conditions of the HP sub-model is then

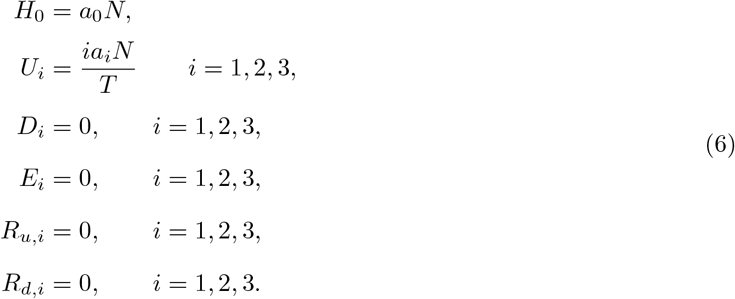

Note that the coefficients *a*_*i*_ are constrained in [0, 1] and must satisfy the system of equations

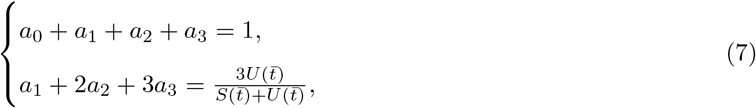

where the first equation comes from the fact that the coefficients *a*_*i*_ are fractions which must sum to 1 and the second equation ensures consistency in the number of undetected people in lockdown. In fact the total number of undetected infected in the lockdown population is given by *H*_1_ + 2*H*_2_ + 3*H*_3_ = (*a*_1_ + 2*a*_2_ + 3*a*_3_)*N* and this number must equal 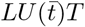. By the definition of *N* and rearranging the terms, the second equation in (7) is obtained.

At the end of the lockdown, the lockdown population mixes again with the free population, thus starting a new free phase, the dynamics of which is described only by the SUDER sub-model. If the switching between lockdown phase and free phase happens at time 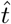, the initial conditions of this sub-model are easily obtained as follows

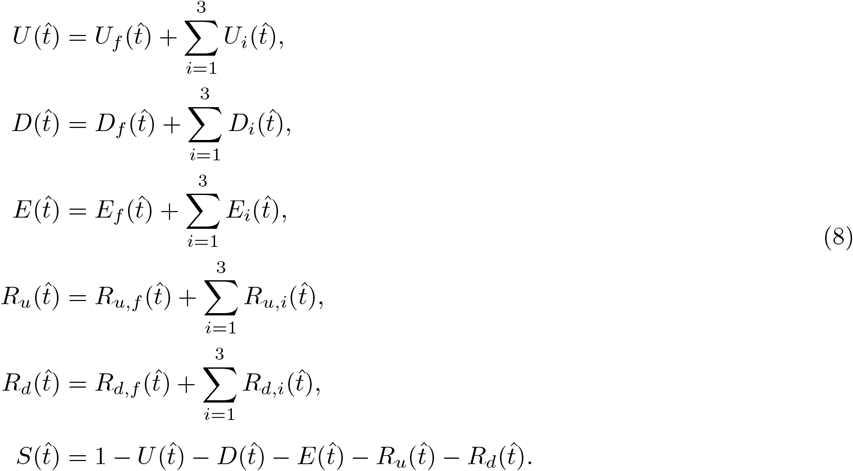

Besides the lockdown percentage, another feature that the model lets the user modify is the *lockdown duration, i*.*e*. the number of days between the lockdown is enforced and lifted. In terms of the mathematical model, the lockdown duration is the amount of time that the FL-Hybrid model spends in the lockdwown phase.

### Discussion on modelling assumptions

Before we discuss the modelling assumptions, it is useful to remark that both the SUDER sub-model and the HP sub-model are mean-field models. This implies that the models themselves do not capture the infection transmission and evolution in a case-by-case fashion, but rather describe the averaged dynamics of the epidemic. In this sense, the parameters of the sub-models are to be meant as average rates over the fraction of population (*i*.*e*. free or lockdown population) described by the sub-models. Carefully choosing the parameters enables making high-precision predictions of the future trends.

In the SUDER and HP sub-models we make the following simplifying assumptions.

- Detected people are properly isolated and do not transmit the infection to susceptible people, *i*.*e*. the number of contacts which detected people have is zero. Therefore, the disease transmission rate from detected infected to susceptible is assumed to be zero. In reality, perfect quarantining does not happen, therefore this rate, although very low, is not zero and depends on a country’s specific policy on detecting and isolating infected individuals. The sub-models might be easily extended to have a non-zero transmission rate from detected infected to susceptible.
- No undetected people will die from the disease, as the development of life-threatening symptoms would lead to a diagnosis before death occurs. This is not always accurate, especially at the beginning of the epidemic when low detection rates and the inability of health services to cope with the high number of cases might lead to official sources under-reporting the number of casualties. The sub-models might be extended by adding an additional compartment for deaths from the undetected stage.
- A recovered person will not become susceptible of re-infections. A prior SARS-Cov-2 infection has been found to be associated to an 83% lower risk of infection^26^, thus justifying this simplifying assumption while still making highly accurate predictions. Re-infections might be introduced in the sub-models by allowing a flow from the *R* compartments to the *S* compartment at a given immunity loss rate.

In the HP sub-model we make a number of additional assumptions aimed at considerably simplifying the intricate household dynamics, yet without compromising the sub-model prediction accuracy.

- The household size is fixed and consists of three members. While in most countries the household size is generally a number between two and four, (3.1 for Israel and 2.1 for Germany in 2019^27^), our HP sub-model requires an integer size. We selected 3 for both countries in order to simplify the theoretical development, but the equations can be easily adapted to have an integer household size as close as possible to a country’s average household size. Note that, although the number of 3 is quite accurate for Israel, yet it also produces accurate results for Germany. Considering households of different sizes is allowed by our model, but it would cause a drastic increase in the number of HP sub-model equations, with minimal benefits in terms of prediction accuracy.
- The coefficients *a*_*i*_ are selected arbitrarily, yet reasonably, in order to satisfy the constraints (7) and the constraint *a*_*i*_ ∈ [0, 1]. Note that since the constraints (7) are a system of two equations in four unknowns, once two of the coefficients, for example *a*_0_ and *a*_1_, are arbitrarily chosen, the remaining ones, *i*.*e. a*_2_ and *a*_3_, are known functions of the first two. Also note that the bounds on *a*_2_ ∈ [0, 1] and *a*_3_ ∈ [0, 1] provide constraints on the values that *a*_0_ and *a*_1_ can assume. The combination of these constraints limits, in fact, the arbitrariness of the choice. For instance, the coefficient *a*_0_ is constrained to a large value in the range [0, 1]. This is consistent with the fact that, as the infected individuals are a small percentage of the population, the vast majority of the households at the beginning of the lockdown will have no undetected members. Additionally, note that the HP sub-model has very low sensitivity over these coefficients. Changing these coefficients does not produce inaccurate predictions as long as the sub-model parameters - inferred from the official data - are suitably updated.
- Within the same household, all undetected infected members are diagnosed at once. This assumption is motivated by the fact that if one member of the household tests positive for the infection, it is very likely that the rest of the household will get tested and diagnosed if infected.
- If infected household members are diagnosed, to avoid spread of the infection the household as a whole will self isolate, as well as its members individually. This prevents new household members from being infected from the outside, and vice versa, and also stops the infection from spreading within the same household.
- All the cases within the same household will evolve in the same way, *i*.*e*. they all either recover or die. Obviously this is not always the case in real circumstances, but, as previously mentioned, this mean-field assumption considerably simplifies the model - making it a simple yet powerful instrument - while still producing good averaged descriptions of the epidemic evolution.
- The households of individuals belonging to the free population are assumed to be part of the free population as well. These include, for instance, the households of key workers. The rationale is that, although the families of the key workers keep isolating at home, they are still virtually in contact with the rest of the free population through the key workers. This explains why official data is compatible with relatively high percentages of free population in the lockdown phase of our model.
- When a susceptible person in an isolating household is infected by an individual outside of the household, it is assumed that this happens only by contact with an infected individual in the free population and not from other households. Introducing also inter-household infections is feasible, but the rate would be very low. Consequently, such a change would just over-complicate the HP sub-model while yielding negligible benefits from the point of view of its dynamic properties.

To conclude this section, we discuss the meaning and practical implications of the lockdown percentage *L* in the lockdown phase. This number, which in our model is defined as the fraction of susceptible and undetected individuals isolating in their own household, should not be considered as an absolute measure of the fraction of population isolating in reality. Instead, once the baseline lockdown percentage is identified by fitting the model with the official data, the model allows the user to assess the outcome of more or less stringent lockdowns by increasing or reducing *L* relative to the baseline.

### Effective reproduction number

In both the free phase and lockdown phase, the epidemic grows if the time derivative of the undetected population is higher than zero. For the free phase, this corresponds to

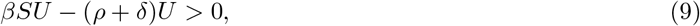

which in turn is equivalent to

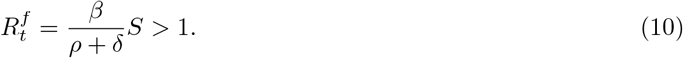

The term 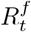 is the effective reproduction number of the epidemic in the free phase provided by the model, while the basic reproduction number is

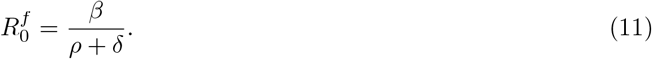

In the lockdown phase the epidemic grows when 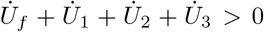. Using the expressions of these derivatives given in (2) and (4) and rearranging we obtain

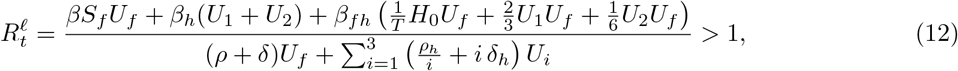

where 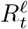 is the effective reproduction number in the lockdown phase. We now want to show that, at the start of the lockdown, the effective reproduction number can be expressed as a function of the lockdown percentage *L*. Let 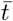 be the time at which the lockdown is enforced and let 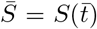 and 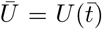 be the portion of susceptible and undetected population at the beginning of the lockdown. Then using the initial conditions 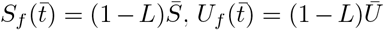, defining *a* = *a*_1_ + 2*a*_2_ + 3*a*_3_ and using equations (5), (6), and (7), the effective reproduction number resulting from the introduction of the lockdown can be expressed as a function of *L* as

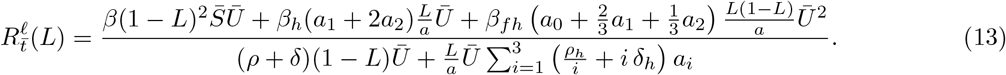

By defining 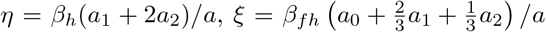 and 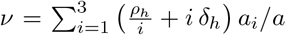, the previous expression reduces to

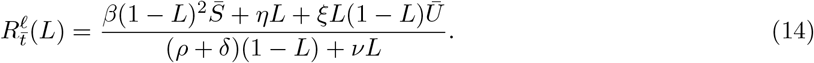

Note that if *L* = 0, *i*.*e*. no lockdown is imposed, the expression of the effective reproduction number in (14) becomes the same as in (10), *i*.*e*. 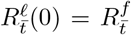 and the effective reproduction number of the free phase is retrieved.

## Data Availability

Official epidemiological data was gathered from the COVID-19 Data Repository by the Center for Systems Science and Engineering (CSSE) at Johns Hopkins University. The repository is available at https://github.com/CSSEGISandData/COVID-19

## Code availability

The code is available at https://github.com/alberto-mellone/FL-Hybrid-model.

## End Notes

### Author Contributions

G.S. conceived the modelling framework, planned and provided oversight for all aspects of the study. Z.G. formulated the initial mathematical model, which was refined by G.S. and A.M. A.M. and Z.G. extracted the data, wrote the code and performed fitting and simulations. Z.G. prepared the figures of the manuscript and A.M. wrote the first draft of the manuscript. All authors contributed to subsequent revisions and provided essential intellectual input into all aspects of this study.

